# Seroprevalence of anti-SARS-CoV-2 IgG antibodies in hospitalized patients at a tertiary referral center in North India

**DOI:** 10.1101/2020.08.22.20179937

**Authors:** Animesh Ray, Komal Singh, Souvick Chattopadhyay, Farha Mehdi, Gaurav Batra, Aakansha Gupta, Ayush Agarwal, M Bhavesh, Shubham Sahni, R Chaithra, Shubham Agarwal, Chitrakshi Nagpal, B H Gagantej, Umang Arora, Kartikeya Kumar Sharma, Ranveer Singh Jadon, Ashish Datt Upadhyay, Neeraj Nischal, Naval K Vikram, Manish Soneja, R M Pandey, Naveet Wig

## Abstract

**Background:** Seroprevalence of IgG antibodies against SARS-CoV-2 is an important tool to estimate the true extent of infection in a population. However, seroprevalence studies have been scarce in South East Asia including India, which, as of now, carries the third largest burden of confirmed cases in the world. The present study aimed to estimate the seroprevalence of anti-SARS-CoV-2 IgG antibody among hospitalized patients at one of the largest government hospital in India.

**Method:** This cross-sectional study, conducted at a tertiary care hospital in North India, recruited consecutive patients who were negative for SARS-CoV-2 by RT-PCR or CB-NAAT. Anti-SARS-CoV-2 IgG antibody levels targeting recombinant spike receptor-binding domain (RBD) protein of SARS CoV-2 were estimated in serum sample by the ELISA method.

**Results:** A total of 212 hospitalized patients were recruited in the study with mean age (±SD) of 41.2 (±15.4) years and 55% male population. Positive serology against SARS CoV-2 was detected in 19.8% patients(95% CI 14.7-25.8). Residency in Delhi conferred a higher frequency of seropositivity 26.5% (95% CI 19.3-34.7) as compared to that of other states 8% (95% CI 3.0-16.4) with p value 0.001. No particular age groups or socio-economic strata showed a higher proportion of seropositivity.

**Conclusion:** Around, one-fifth of hospitalized patients, who were not diagnosed with COVID-19 before, demonstrated seropositivity against SARS-CoV-2. While there was no significant difference in the different age groups and socio-economic classes; residence in Delhi was associated with increased risk (relative risk of 3.62, 95% CI 1.59-8.21)

## Introduction

Since the first novel Corona virus disease 2019 (COVID-19) case in India on January 30, 2020(1), the number of cases has steadily increased and has crossed 2.9 million by 21st August 2020(2). COVID-19 frequently results in asymptomatic and mild infections, which may remain undetected, and the actual number of cases is a matter of speculation. In a country with a population above 1.35 billion, ~20,000 tests per million were being performed as of mid August (3), suggesting that a large cohort of cases, who are either asymptomatic or of mild severity, remain undetected. Seroprevalence studies reflect the proportion of people exposed to the infection, and find utility in accounting for the true burden of infection in the community. Seroprevalence studies for SARS-CoV-2 have been conducted in different settings, including community(4)(5)(6)(7), special populations like parturients (8), liver disease patients(9), hemodialysis patients(10), blood donors(11), health care workers(12)(13) as well in hospitals (14). However, few seroprevalence studies have been reported from India. A recent sero-surveillance study in Delhi conducted between 27^th^ June 2020 to 10^th^ July 2020, done by National Centre for Diseases Control(NCDC)in collaboration with Government of National Capital Territory of Delhi, found 23.48% of the population of Delhi to be seropositive for SARS-CoV-2 IgG antibody by ELISA(15). The second serological survey conducted between 1^st^-7^th^ August found the figure increased to 28.3%. This suggests that a significant proportion of Delhi’s population may have been exposed to the virus, much more than the number of cases(detected by RT-PCR, CB-NAAT, antigen tests etc) indicate. However, no hospital based seroprevalence study has yet been reported from India. A tertiary care hospital caters to a heterogeneous patient profile, with multiple comorbidities, who also pose a higher risk of developing complications from COVID-19. Seroprevalence among admitted patients in a hospital provides an estimate of seroprevalence in the community, and indirectly tells us about the spread of infection in that community. In this study, the seroprevalence of SARS-CoV-2 IgG antibody in patients admitted to a major tertiary hospital in Delhi was estimated as well as characteristics of the seropositive patients vis-à-vis the seronegative patients were determined.

## Material and Methods

This was a cross-sectional study conducted at All India Institute of Medical Sciences (AIIMS), New Delhi between 9^th^ June and 8^th^ August 2020,among patients who were admitted to the medicine wards and intensive care unit (ICU). Our tertiary care hospital, situated in in the heart of national capital of India, caters to patients from most states of North India. However, due to the COVID-19 pandemic-related travel restrictions, majority of admissions during the study period were of patients hailing from Delhi, with few from surrounding states of Haryana and Uttar Pradesh. Consecutive patients(age >14years) admitted to medicine ward/ICU, whose reverse transcriptase-polymerase chain reaction (RT-PCR)/ Cartridge based Nucleic acid amplification test (CB-NAAT) for SARS-CoV-2 from nasopharyngeal/nasal swab and/or oropharyngeal swab were negative. The workflow of the study is depicted in Figure 1. Cases with confirmed COVID-19 in the past were not included in the primary analysis but was used to compute the corrected seroprevalence estimate. After obtaining written consent, 4 ml of blood was drawn, serum was stored at −80°C and was defrosted before testing. Anti-SARS-CoV-2 IgG antibody was detected using ELISA method, developed and validated “in-house” at the Translational Health Science and Technology Institute (THSTI), Faridabad (Mehdi et al, manuscript under preparation). The ELISA targeted IgG antibody against recombinant spike receptor-binding domain (RBD) protein of SARS CoV-2(16). The estimated sensitivity of THSTI’s IgG ELISA for RT-PCR positive cases was 88.24% (95% CI: 82.05% to 92.88%) in individuals after 20 or more days from onset of symptoms or from a positive RT-PCR test; and specificity was 99.79% (95% CI: 98.82% to 99.99%) as determined from the pre-pandemic serum samples (Mehdi et al, manuscript under preparation). It was decided to randomly select atleast 60% of the samples positive in RBD ELISA and test them using Euroimmun IgG ELISA for SARS CoV-2 (17) and Zydus Kavach IgG ELISA for SARS CoV-2.(18) Patients details including demographic features and relevant details of present hospitalization were collected using a case record form filled by trained personnel.

**Figure 1.**
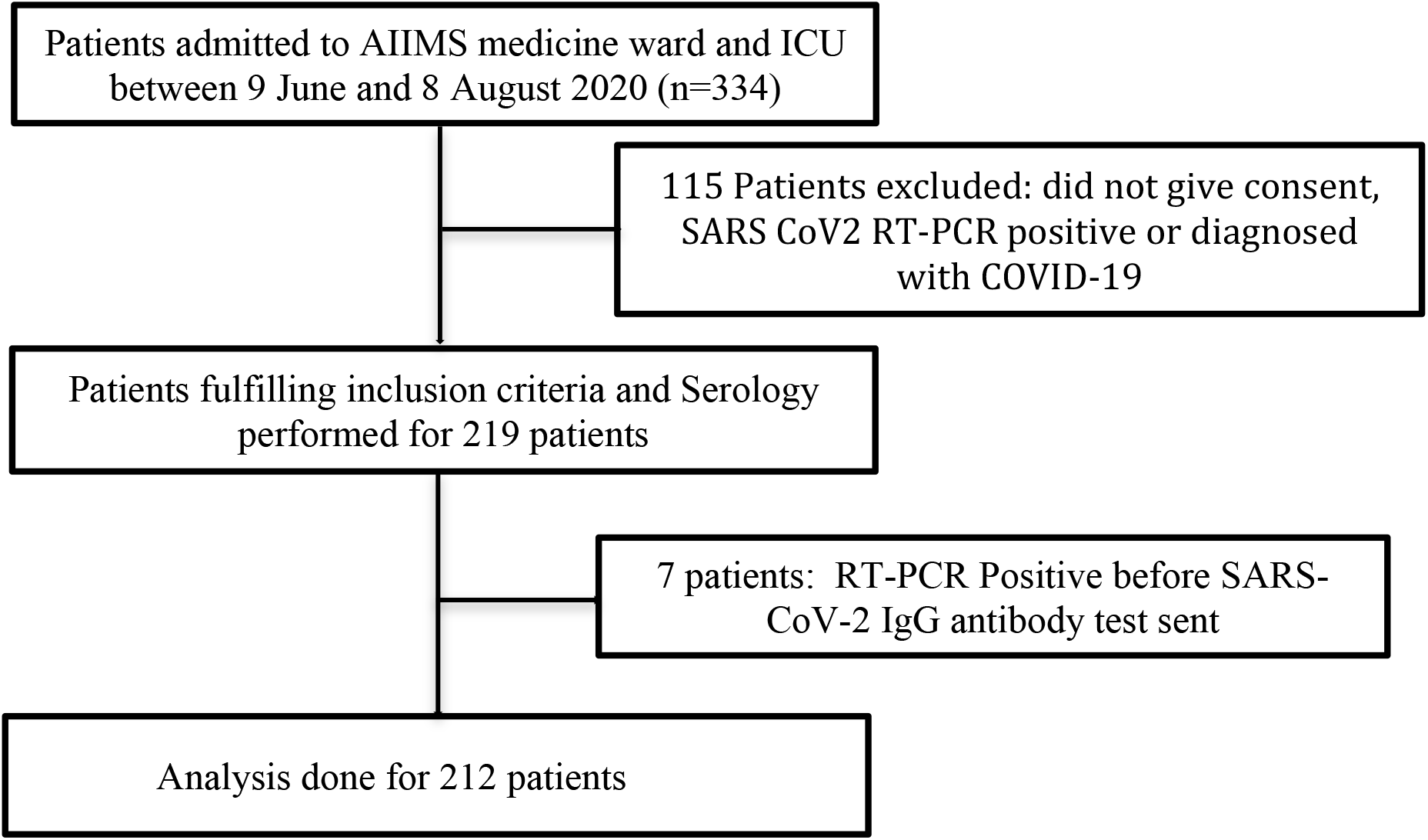

To estimate the sample size, we performed a pilot study on 50 subjects, 6 of whom were positive for the IgG ELISA. Thus, assuming the sero-prevalence of COVID-19 to be 12% (n=6/50) with precision of 5% and a non-response of 10%, the estimated sample size was 180.

## Statistical Analysis

STATA Version 12.1 (StataCorp) was used for the statistical analysis. Categorical variables were calculated as frequency and percentage, continuous variables were represented as mean (± SD). Chi-square test and t test were used to calculate the statistical differences between categorical variables and continuous variables respectively. A p-value of <0.05 was considered as statistically significant.

## Results

Between 9^th^ June 2020 to 8^th^ August 2020, 212 patients were recruited for the study, 117 (55%) of whom were male. The mean age(±SD) of the patients was 41.2 (±15.4) years. Socioeconomic status of the admitted patients was quantified using the modified Kuppuswamy scale(19). Majority (84.4%) of the patients belonged to socioeconomic class III (middle) to class V (lower). At least one comorbidity, such as hypertension, diabetes mellitus, chronic kidney disease, chronic lung disease, heart disease, human immunodeficiency virus (HIV) infection, past or recently diagnosed tuberculosis, was documented in 153 (72%) patients. Various characteristics of the patients are highlighted in Table 1. At the time of presentation, 107(50.5%) patients were admitted with clinical symptoms suggestive of infective etiology. No statistically significant difference in seroprevalence of anti-SARS-CoV-2 IgG antibodies were observed between patients admitted with or without comorbidities. (Refer Table 1 in Appendix). In this study, 59% of patients had a BCG scar, or gave a history of BCG vaccination, but BCG vaccination was found to have no significant effect on the prevalence of anti-SARS-CoV-2 IgG seropositivity.

**Table 1:** Demographic profile and baseline characteristic of study population

| Variables | Total<br>(n=212) | SARS-CoV-2<br>IgG antibody<br>positive(n=42) | SARS-CoV-2<br>IgG antibody<br>Negative(n=170) | p Value |
| --- | --- | --- | --- | --- |
| Age (Mean±S.D) | 41.2±15.4 | 42.9±13.9 | 40.8±15.8 | 0.43 |
| Males | 117 (55%) | 23 (55%) | 94 (55%) | 0.95 |
| Any comorbidity | 153 (72%) | 31 (74%) | 122 (72%) | 0.79 |
| BCG vaccination | 125 (59%) | 20 (48%) | 105 (62%) | 0.09 |
| Addiction | 49 (23%) | 9 (21%) | 40 (24%) | 0.77 |
| Residence (Delhi) | 136<br>(64.2%) | 36 (86%) | 100 (59%) | 0.001 |
| Outside Delhi | 76 (35.8) | 6 (14%) | 70 (41%) |  |
| Socio-economic status (Modified Kuppuswamy scale)* |  |  |  |  |
| Upper | 2 (1%) | 0 (0%) | 2 (1%) | 0.70 |
| Upper middle | 28 (13%) | 5 (12%) | 23 (14%) |  |
| Middle | 72 (34%) | 11 (27%) | 61 (36%) |  |
| Upper lower | 82 (39%) | 19 (46%) | 63 (37%) |  |
| Lower | 25 (12%) | 6 (15%) | 19 (11%) |  |

\*Data missing for 3 patients, p value calculated using Fischer exact test

25 positive samples on RBD ELISA were retested on Euroimmun ELISA and Zydus Kavach IgG ELISA. 23 out of 25(92%)were positive on Euroimmun ELISA, 17 out of 25(68%) were positive on Kavach ELISA. Only one sample was positive on RBD ELISA but negative on Euroimmun and Kavach IgG ELISA. The overall prevalence of anti-SARS-CoV-2 antibody among admitted patients was 19.8% (95% CI 14.7-25.8). Seven patients, whose SARS-CoV-2 RT-PCR was found to be positive predating collection of samples for SARS CoV-2 IgG antibody, when added to the analysis yielded the corrected prevalence of 22.4%(95% CI 17.0-28.5). One hundred thirty six(64.2%) patients were continuously residing in Delhi since before the pandemic began, of which, 36 (26.5%) (95% CI 19.3-34.7%)tested positive against SARS-CoV-2 IgG. The corrected seroprevalence of patients from Delhi (after adding results of 5 out of 7 patients), after considering SARS-CoV2 RT-PCR/CB-NAAT positive patients increased to 29.1%(95% CI 21.7-37.3). Around 76(35.8%) patients resided in outside states like Haryana, Bihar, Uttar Pradesh, of which, 6(8%) (95% CI 3.0-16.4.) tested positive for the SARS-CoV-2 IgG antibody, the difference being statistically significant with p value 0.001. Only 1 (2.4%) (95% CI 0.06-12.6). SARS-CoV-2 seropositive patient gave history of Influenza like illness(ILI) symptoms in past 6 months. In patients who were seropositive against anti-SARS-CoV-2 antibody, no significant differences in history of household contact with confirmed COVID-19 cases or of travel in public vehicles were noted compared to seronegative cases.

In 1^st^ week, calculated seroprevalence among admitted cases was 32%, which decreased in subsequent next 2 weeks to 11%. In week 4 and 5, seroprevalence was 18% and 25% respectively; in week 6, only 14% patients were positive for SARS-CoV-2 IgG antibody, 0 cases were reported in week 7, and in week 8 again seroprevalence rose to 27%(Refer Table 2 in Appendix). Week 2 was taken as reference week for calculation of relative risk, as positivity rate was not in extreme range i.e. neither too high nor too low, and number of participants in this group was similar when compared with other age groups.(For details refer table 2 in Appendix)

**Table 2:**
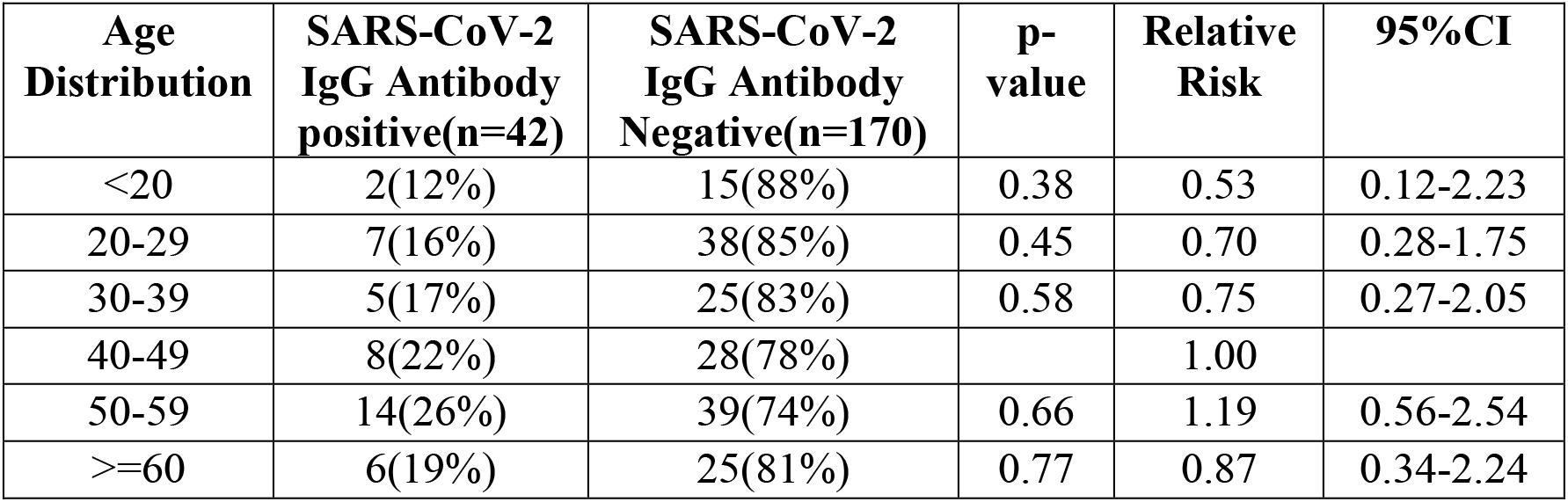
Age-wise distribution of SARS-CoV-2 IgG antibody positive case (Age group 40-49 taken as reference)

Seroprevalence of COVID-19 was highest between 40 to 59 years and was calculated to be 22% and 26% for 40-49 and 50-59-year age group respectively. It was approximately 15%, between 20 to 39 and 19% for patients who were 60 years of age and above(Table 2). Age group 40-49 years was taken as reference group to calculate the relative risk across all age groups. No statistically significant difference among antibody positivity rate was noted in any age group.

## Discussion

This hospital based seroprevalence study in the national capital, showed that 19.8 % of patients, who do not have a confirmed laboratory diagnosis of COVID-19, either in the past or at the time of presentation, had IgG antibody to SARS-CoV2. As the population of the study were largely unselected and can be assumed to represent the population of the catchment area of the hospital, this study indicates that a substantial proportion of the population of Delhi have been exposed to the SARS-CoV-2 after the onset of the pandemic in December 2019. A comparison of the age-wise distribution of the study population showed that across three out of six age-groups, the study population was statistically similar to population of Delhi (as per 2011 census data)(20). (See appendix Table 3)

The implications of this finding are manifold. First, it implies that every 1 out of 5 patients presenting to a hospital without a prior history of confirmed COVID-19 have had exposure to anti-SARS-CoV-2. Moreover, taking in account the corrected prevalence of 29.1 *%* among patients residing in Delhi, the estimate of prevalence of anti-SARS-CoV-2 IgG antibodies in the population of Delhi appeared to be ~3 out of 10. Since majority (>97%) of people have had minor or no symptoms attributable to COVID-19, it signifies that the viral exposure have lead to predominantly asymptomatic infection. Incidentally, a recent sero-survey in Delhi (15) pegged the seroprevalence of COVID-19 in Delhi population at 23.48% (samples collected in late June and early July) and 28.3%(samples collected in first week of august); which also supports the finding of our study. Second, the seroprevalence data also reflects on the Infection fatality risk (IFR) of COVID-19. Since, it is not possible to diagnose all symptomatic infections as well as asymptomatic infections, the IFR of SARS-CoV2 has been difficult to ascertain and has been a matter of intense debate(21). Sero-surveys are thought to be invaluable in diagnosing the proportion of infected individuals irrespective of symptoms. The data from this sero-survey indicate that the IFR of SARS-CoV2, is around 0.084% (4082 deaths were reported in Delhi till 8^th^ Aug 2020; population of Delhi was taken to be 18710922(22), which is much lower than that previously estimated(24). It would be worthwhile to remember that during the initial period of the pandemic, Crude Fatality Rate (CFR) as suggested by WHO was around 3.4%(23).However, subsequently the mortality rate has been revised several times and appears to shrink further in the light of the results of this study. Very low IFR has also been inferred from seroprevalence studies in other Asian countries including Iran(24), China(with the exception of Wuhan)(25), and Israel(26). The country-wide seroprevalence estimate done in India which was initiated in May 2020 had showed a country wide prevalence of infection at 0.73% with an implied IFR of 0.08%(27). A few reasons for lower IFR in Asian countries including India has been suggested including previous exposure to coronaviruses, genetic differences and lower infectious load.(28)

The seroprevalence of patients coming from Delhi was significantly higher than that from outside Delhi indicating that residence in Delhi was a significant risk factor for having anti-SARS-CoV-2 seropositivity among admitted cases. This seems to reflect not only the total number of detected cases in the respective states, but also the relative percentage of state population infected. The true extent of spread of infection and its surrogate seroprevalence is expected to be proportional to the percentage of population diagnosed as COVID-19 (by RT-PCR, CB-NAAT etc) divided by the number of tests conducted(29). Figure 2 shows the total number of cases in the states of Delhi, Uttar Pradesh, Bihar and Haryana on June 9^th^ and August 8^th^.(30) Table 3 in Appendix shows the relative population percentage (of India) (31) and Figure 3 shows for the three states of Delhi, Haryana and Uttar Pradesh, the seropositivity percentage from the present study along with contribution to country’s population in %, contribution to total cases in India in %, ratio of positive cases to country* 10 / % of country population*tests per million (ratio). As it is apparent from the graph, the seropositivity percentage of patients from Delhi was higher than that from adjacent states of Uttar Pradesh and Haryana and seemed to track the ratio mentioned above.

**Figure 2:**
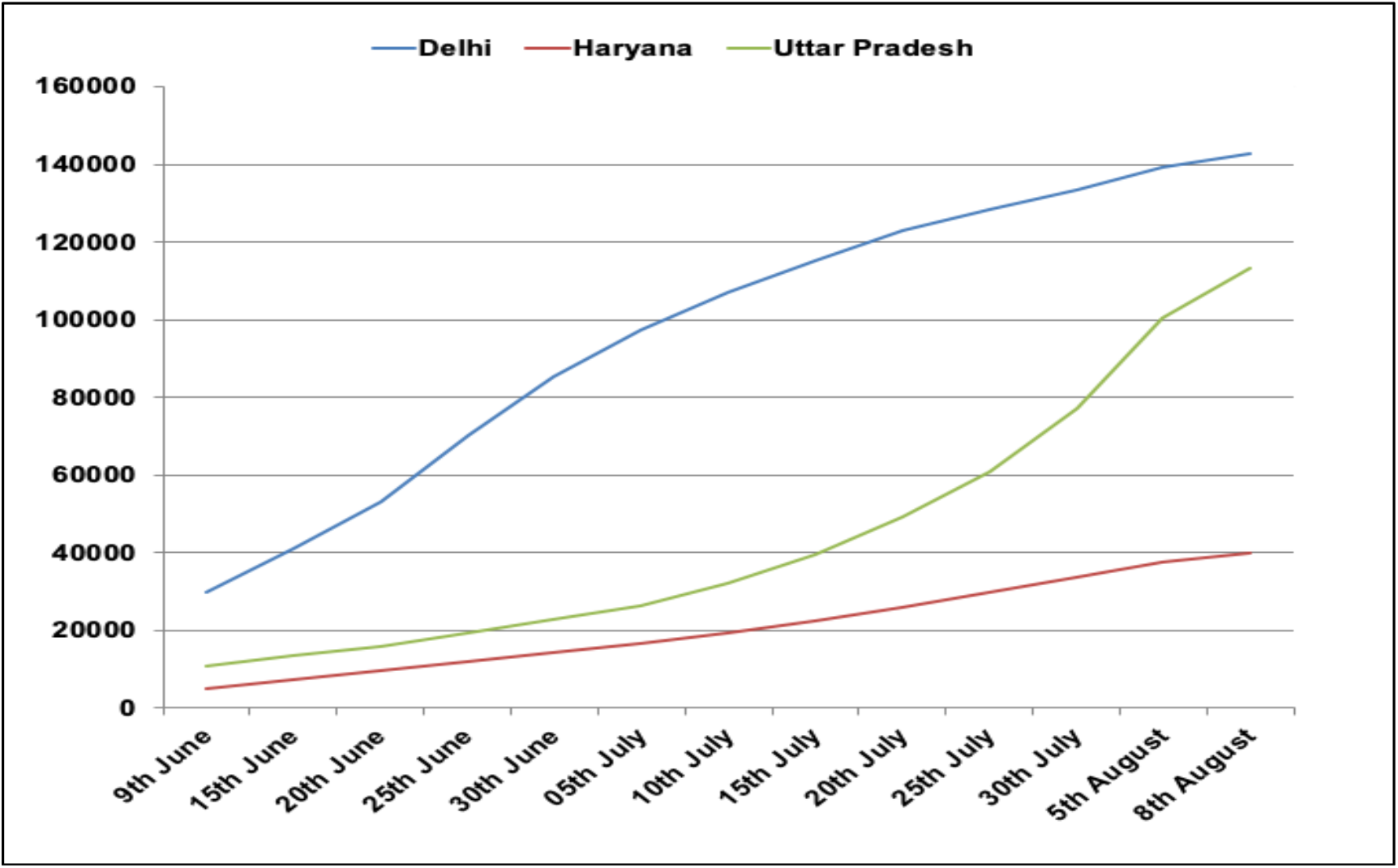
Number of COVID-19 positive cases – State wise data(prsindia.org/covid-19/cases)

**Figure 3:**
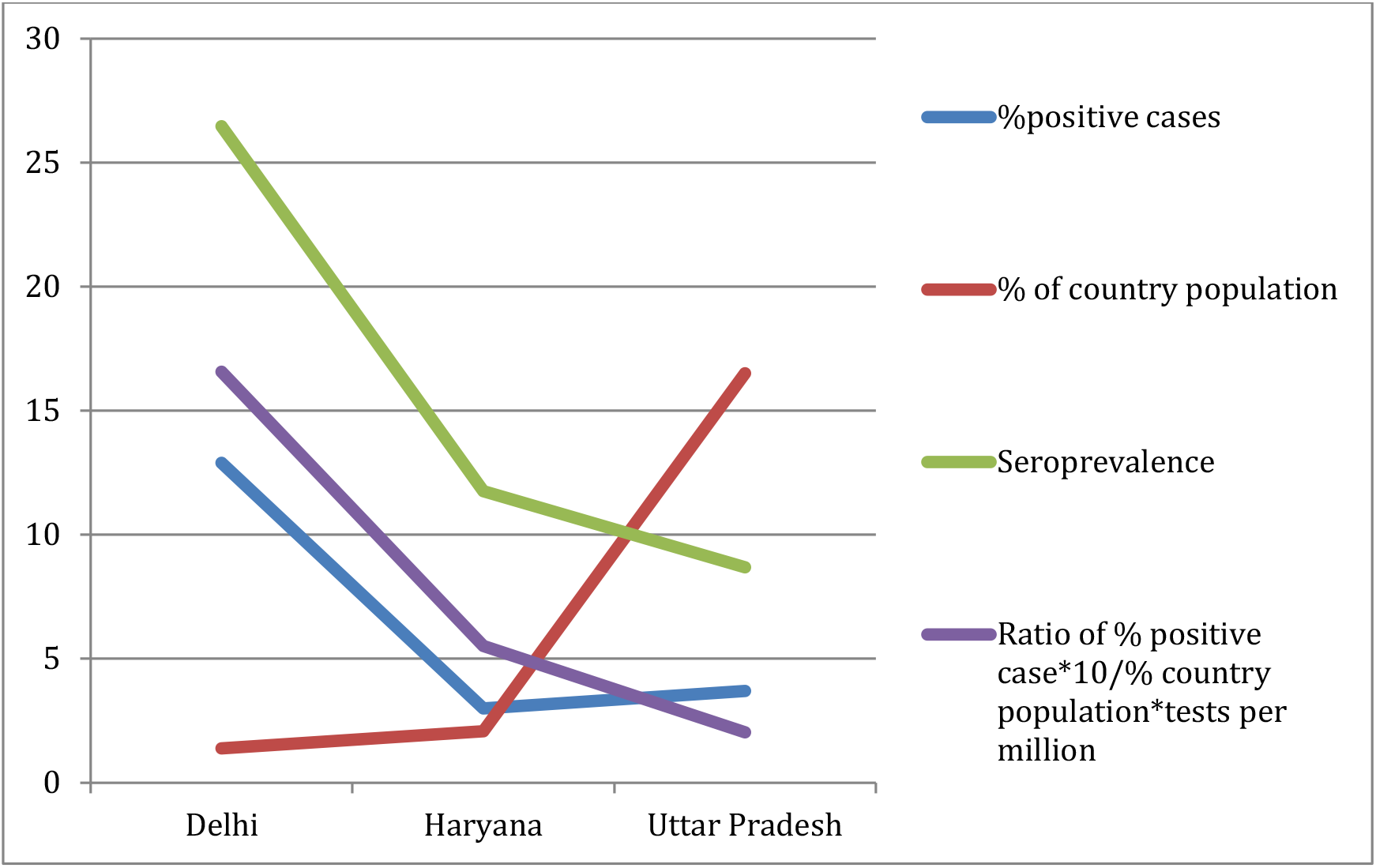
State wise data showing seroprevalence, percentage positive cases, ratio of positive case to country population*test per million(data as on 8^th^ August), prsindia.org/covid-19/cases

None of the age groups as well as socio-economic classes was found to have a higher relative risk of infection suggesting that the exposure to infection was not confined to any particular group. It has been suggested that BCG vaccine has a protective role against Covid-19 in countries with high TB burden, where universal BCG vaccination at birth is a part of national policy. It has been observed that countries with universal BCG vaccination have a low number of COVID-19 cases(32) and lesser severity(33), however, whether it is a chance occurrence, is still unknown and need to be verified by well-conducted studies(34). In our study, prevalence of BCG Scar or vaccination was 59% This low prevalence can be attributed to recall bias, fading of BCG scar (35) and also due to low vaccination rate before universal immunization was started (1985). Our study did not find any association between BCG vaccination and presence of seropositivity of COVID-19 Antibody.

The strengths of the present study include, that this is the first such seroprevalence study from a hospital in India and confirmation of seropositivity by ELISA tests (>60% positive results corroborated by two other established ELISA kits). The study also had some inherent weaknesses. As it was a hospital-based study, the population in the study cohort may not have been truly representative of the population at large. However, the concordance of the seropositivity data with that of the recently conducted sero-surveillance (15) lays credence to the validity of the results. Also since anti-SARS-CoV-2 antibody levels is known to wane over few months, some “exposed” cases might have inadvertently missed in this cross sectional study.

## Conclusion

The present study indicates that around 19.8% of cases admitted to hospital, without prior history of confirmed COVID-19, had been exposed to SARS CoV-2 causing predominantly mild/asymptomatic infections. Residency in Delhi conferred a higher frequency of antibody positivity (26.5%) as compared to that of other states(Uttar Pradesh/Haryana) (8%). No particular age groups, sex or socio-economic class showed higher relative risk for seropositivity. The calculated IFR of COVID-19 in Delhi seems to be a much less (0.08%)than that previously expected.

## Data Availability

Data will be provided on request

## Notes

### Competing Interest Statement

The authors have declared no competing interest.

### Funding Statement

This work is supported by intramural funds of All India Institute Of Medical sciences, New Delhi. No external funding was received. Special acknowledgment to Translational Health Science And Technology Institute for processing SARS-CoV2 serum sample for antibody detection

### Author Declarations

INSTITUTE ETHICS COMMITTEE FOR POST GRADUATE RESEARCH, ALL INDIA INSTITUTE OF MEDICAL SCIENCES, NEW DELHI, 110029,INDIA

